# Cytocapsular Tube-Based Precise Cancer Diagnosis in Colon Cancers

**DOI:** 10.1101/2021.07.28.21261279

**Authors:** Tingfang Yi, Gerhard Wagner, Stephen Ganshirt

## Abstract

Colorectal (colon) cancer ranks second in terms of cancer lethality worldwide. It is estimated that approximately 1.93 million new colorectal (colon) cancer cases and approximate one million colorectal cancer deaths occur worldwide in 2020 alone. Cancer metastasis is a major source of cancer lethality. Mechanisms underlying colorectal cancer metastases are long-term obscure. Recently, a new organelle associated with aggressive cancer cells, dubbed cytocapsular tube, was recognized to conduct cancer cell metastases in tissues *in vivo*. Here, we investigated the roles of cytocapsular tubes in colon cancer metastasis in 9 subtypes of colon cancers with original clinical colon cancer tissues, paracancer tissues of colon cancer, and metastatic colon cancers in multiple other tissues and organs. We found that colon cancer metastasized via cytocapsular tubes and their networks in clinical tissues *in vivo*. Furthermore, we applied the cytocapsular tube analyses in precise cancer diagnosis in clinical colon cancers and obtained better outcomes in clinical colon cancer management.

## Introduction

Colorectal (colon) cancer is among the top 3 most aggressive and popular cancers globally^1^. It is estimated that approximate 1.93 million new colon cancer cases and approximate one million colon cancer deaths occurred worldwide in 2020 alone^1^. Overall, colorectal ranks third in terms of incidence, but second in terms of mortality. The precise and verifiable prognosis and diagnosis of colon cancer is a prerequisite for the timely, appropriate and effective therapy, treatment and management of suspicious, progressive, symptomatic, metastatic and recurrent colon cancer ^2-3^. However, the absence of effective prognosis methods, misdiagnosis, pseudo negative, pseudo positive, and difficulty to indentify or classify in clinical operation, which are caused by low resolution at mm or cm levels of conventional diagnosis methods and technologies, have negatively affected the therapy and treatment outcomes of colon cancer^3-4^. Sporadic cancers, surface skin cancers, and all kinds of polyps, which have sufficient spaces for uncontrolled cancer cells to extend and do not show abnormal high cell density, cannot be detected by high cell density based diagnosis methods, such as computer tomography scan, enhanced computer tomography scan, or positron emission tomography–computed tomography scan (dense radioactive sugar fluorodeoxyglucose-18 of abnormally dense cells). Effective checks and evaluation methods of colon polyps are generally lacking because: 1) the colon polyps have sufficient space to extend in the colon lumen and do not show high cancer cell density threshold needed for classical diagnosis detection, 2) there are no reliable gene/protein molecular markers and derived antibodies for cancer diagnosis on colon polyps with immunochemistry staining. For Stage II colon cancer patients with aggressive surgery, there is a clinical dilemma whether to follow up with chemotherapy due to the debatable outcomes. Reliable methods are absent for precise identification of early colon cancerous transformation at the μm level (including colon polyps)^5-6^.

Therefore, there is an urgent need for technologies that can verifiably identify colon cancer metastases and engage biomarkers that effectively detect cancer metastasis pathways with high sensitivity and specificity. Colon cancer-cell dissemination is a major source of colon cancer lethality^7-8^. Mechanisms underlying colorectal cancer metastases are obscure. More recently, cytocapsular tubes were recognized as new organelles for universal solid cancer cell metastases in tissues *in vivo*^*9*-10^.

Here, we investigated the predictive/diagnostic roles of cytocapsular tube biomarkers in 9 subtypes of original clinical colon cancers, paracancer tissues of colon cancer, metastatic colon cancers in multiple other tissues and organs. We found that cancer metastases of 9 subtypes of colon cancers can be mapped with antibody staining against cytocapsular tube biomarkers in clinical tissues *in vivo*. Furthermore, we applied the cytocapsular tube analyses in clinical colon cancer diagnosis and obtained better outcomes in clinical colon cancer management.

## Methods

### Patients and Sample Collection

The patients providing formalin-fixed paraffin-embedded tissue samples gave informed consent that they understood that the biopsies (needle biopsy or surgical biopsy, from US Biomax) were performed for in vitro research purposes only. Comparative deidentified samples of normal tissues, benign tissues, carcinoma *in situ*, cancer, paracancer, metastatic tissues with their cancer stages identified according to the tumor (T), node (N), and metastasis (M) TNM system (cancer stages: 0, I, II, III, IV) were obtained from archival materials. The cancer, paracancer and metastatic cancer tissues, in which the original cancer niches were identified by indicated cancer specific molecular markers, were identified by hospital pathology laboratories and obtained from archival material.

### Histology and Immunohistochemical testing

Cancer lesion samples were obtained from the index patients from clinical hospitals in USA (356 specimens from US Biomax; others samples directly from clinical hospitals). Samples were processed for histopathological evaluation with the use of hematoxylin and eosin staining. Immunohistochemical fluorescence tests were performed to stain cytocapsular tubes using rabbit anti-CM-01 monoclonal and polyclonal primary antibodies (self-made), mouse anti-gamma-actin monoclonal primary antibodies (Abcam, ab123034), Goat anti-Mouse IgG (H+L) Highly Cross-Adsorbed Secondary Antibody, Alexa Fluor Plus 555 (Thermo Fisher), and Goat anti-Rabbit IgG (H+L) Highly Cross-Adsorbed Secondary Antibody, Alexa Fluor Plus 488, Thermo Fisher).

### Data collection

Cytocapsular tubes (CTs, not sectioned, longitudinally sectioned, and cross sectioned) without degradation (3∼6μm in measured diameter) were counted using a fluorescence microscope and ImageJ. The presence of CTs degrading into thick strands (1∼2μm in measured diameter), thin strands (0.2∼1μm in measured diameter), or the disintegration state were reported without quantification.

### Statistical analysis

For each specimen, the number of fully intact cytocapsular tubes was counted in 5 areas (0.35mm x 0.35mm, length x width) of the sample (top, bottom, left, right, and center), and the cytocapsular tube density (CT/mm^2^) was calculated and determined for each area. The average CT density across the 5 sites was treated as the specimen’s overall CT density.

## Results

### Cytocapsular tubes are universally present in original niches of 9 subtypes of colon cancers

Using immunohistochemistry (IHC) staining assays with anti-CM-01 and anti-gamma-actin primary antibodies, and high resolution fluorescence microscopes, we found that there are many cytocapsular tubes (CTs) in diverse lifecycle stages in the original colon tumor blocks (**Fig.1A**). Long, curved and entangled colon cancer cell CTs form superstructures in compact or sporadic colon cancer tissues. There are many colon cancer-cell CTs degraded into very thin strands. Colon cancer cell CTs in diverse stages in CT lifecycle coexist. Colon cancer cells migrate in colon cancer CTs in tissues *in vivo*. The CT masses form irregular super-large and complex structures in the compact colon cancer tissues (**Fig.1A**).

**Figure 1.**
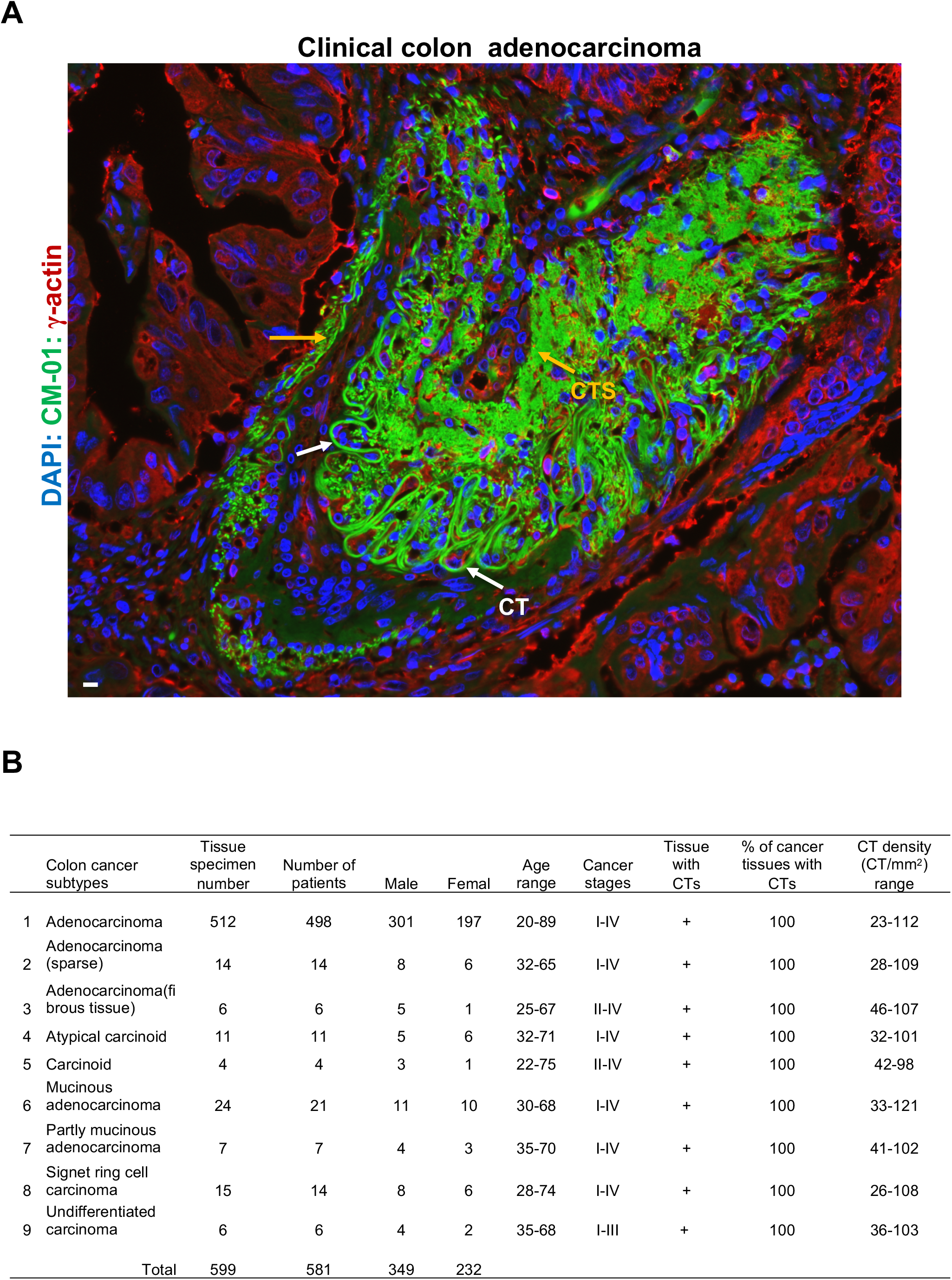
Cytocapsular tubes universally present in clinical colon cancer original niches. **(A)** Fluorescence microscope image of clinical colon adenocarcinoma. Sample is stained with DAPI (blue), anti-CM-01 antibody (green), and anti-gamma actin antibodies. There are massive curved cytocapsular tubes (merged as golden color, CTs, white arrows), entangled together and form superstructures in the compact colon cancer original niche tumor block. Many CTs degrade into thin CT strands (CTS, orange arrows). Scale bar,10μm. **(B)** Quantitative analyses of CTs in 9 subtypes of clinical colon cancers.

Next, we investigated CTs in original niches of 9 subtypes of colon cancers: adenocarcinoma, adenocarcinoma (sparse), adenocarcinoma (fibrous tissue), atypical carcinoid, carcinoid, mucinous adenocarcinoma, partly mucinous adenocarcinoma, signet ring-cell carcinoma, and undifferentiated carcinoma. These 599 colon cancer tissue samples were from 581 colon cancer patients (349 male and 232 female) with various cancer stages (I-IV), male and female, and broad age range (20-89 years old). Cytocapsular tubes universally present in all the examined 599 colon cancer samples (100% colon cancer tissues harbor CTs) with diverse CT densities (from 23CT/mm^2^ to 121CT/mm^2^, **Fig.1B**). Colon cancer cells migrate in CTs and networks in original niches of colon cancers in all the checked 599 colon cancer -tissue samples.

### Paracancer tissues of 9 subtypes of colon cancers harbor high density colon cancer cell cytocapsular tubes for colon cancer cell dissemination

Paracancer tissues, conventionally named “normal adjacent tissue (NAT)”, are the sites that cancer-cell CTs pass through to reach neighboring or far-distance tissues and organs. We examined paracancer tissues in colon cancer, and observed numerous curved CTs form highly compact CT masses in all the checked locations, suggesting that these paracancer tissues are pseudo-negative sites that harbor many CTs, which provide membrane enclosed physical pathways for colon cancer cell dissemination in multiple directions (**Fig 2A**).

**Figure 2.**
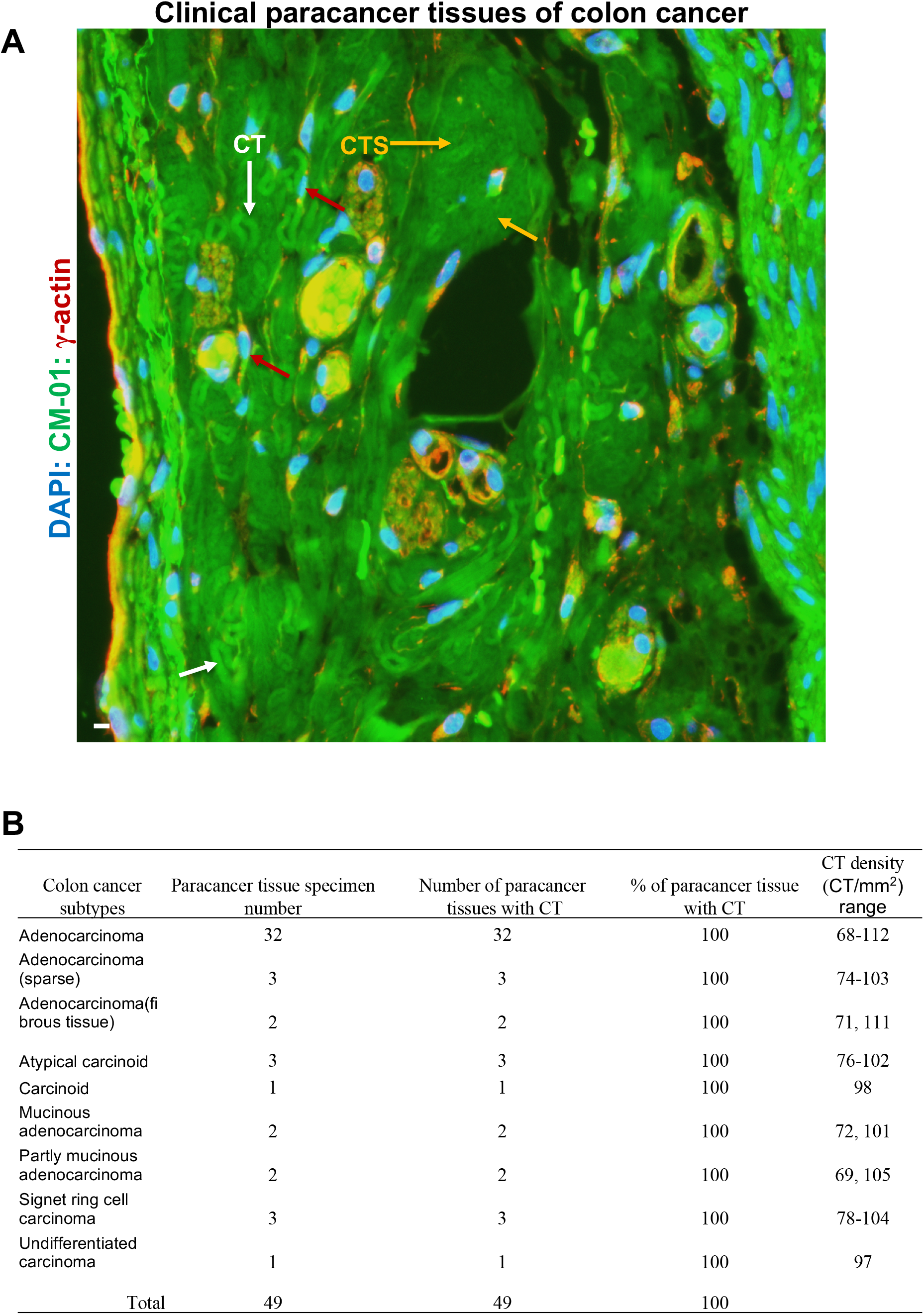
Cytocapsular tubes are widely distributed in paracancer tissues of clinical colon cancers and cancer cells migrate in CTs in metastasis. **(A)** Fluorescence microscope image of paracancer tissues of clinical colon adenocarcinoma. There are a large quantities of highly curved cytocapsular tubes (CTs, white arrows) piled together and CT masses in the paracancer tissues in the colon. Many CTs degrade in to thin CT strands (CTS, orange arrows). There are many colon cancer cells (red arrows) migrating in CTs. **(B)**Quantitative analyses of CTs in the paracancer tissues of 9 subtypes of clinical colon cancers.

Next, we investigated the paracancer tissues of 9 subtypes of colon cancers: adenocarcinoma, adenocarcinoma (sparse), adenocarcinoma(fibrous tissue), atypical carcinoid, carcinoid, mucinous adenocarcinoma, partly mucinous adenocarcinoma, signet ring cell carcinoma, and undifferentiated carcinoma (n= 1-32 samples per subtype). In the 49 examined paracancer tissues of these 9 subtypes of colon cancers, we found that 100% paracancer tissues in all the checked 9 subtypes of colons display numerous CTs with high CT density (68-112 CT/mm^2^, **Fig.2B**) in all the examined colon cancer paracancer tissues. The average CT densities in paracancer tissues in colon cancers are statistically higher that that of original niche in colon cancers (**Fig. 1B and 2B**). These observations evidenced that paracancer tissues variably harbor plenty of CTs and CT networks providing membrane-enclosed freeways for cancer cell metastasis and dissemination.

### Metastatic colon cancer cells disseminate via cytocapsular tubes in cross tissue and organ colon cancer metastasis

Cross-tissue and organ-cancer metastases are major sources of cancer spreading into multiple organs, leading to biological function failure of these organs and are followed by deaths of cancer patients. We analyzed CT spread of metastatic colon cancers to the breast and found that there are massive amounts of colon cancer-cell CTs in breast tissues at diverse stages (with or without degradation into thin strands, **Fig.3A**). The observed CTs form irregular morphologies exhibiting super-large structures, and there are many cancer cells migrating in colon CTs in the breast tissues (**Fig. 3A**).

**Figure 3.**
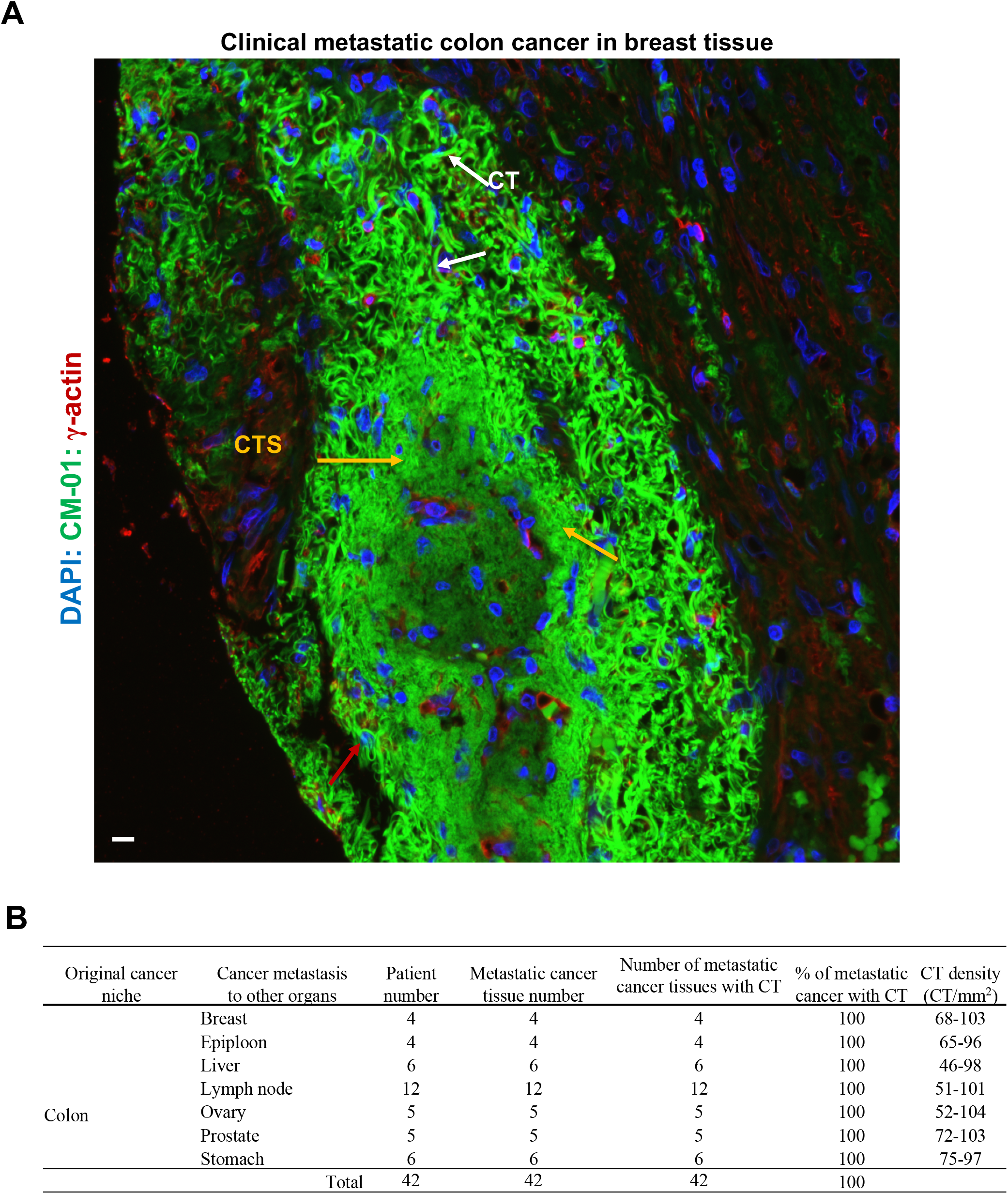
Cytocapsular tubes and networks conduct cross-organ colon cancer cell metastasis. **(A)** Fluorescence microscope image of metastatic colon cancer in breast tissue. There are many highly curved colon cancer cell cytocapsular tubes (CTs, white arrows) that form CT masses in the breast tissues. Some colon cancer cell CTs degrade into thin CT strands (CTS, orange arrows). Colon cancer cells (red arrows) are in migration in CTs in breast tissues. **(B)** Quantitative analyses of colon cancer cell CTs in multiple metastatic tissues in neighboring and far-distance organs.

Next, we examined colon cancer cell CTs in multiple metastatic colon cancers, from colon metastasis to breast, epiploon, liver, lymph nodes, ovary, prostate and stomach. We found that CTs universally (100% CTs in metastatic cancers) present in all these metastatic tissues/organs with relatively high CT densities of 46-104 CT/mm^2^ (**Fig. 3B**). There are many colon cancer cells in migration in colon cancer cell CTs in the metastatic organs. These observations evidenced that colon cancer CTs conduct colon cancer cells metastasis crossing neighboring and far-distance tissues and organs.

In summary, colon cancer cell CTs universally present in original niches of all checked 9 subtypes of colon cancers, in paracancer tissues of all examined 9 subtypes of colon cancers, and in 7 different metastatic colon cancers reached in far-distance organs. Cytocapsular tubes and networks are membrane-enclosed physical freeway systems for cancer cell metastasis to neighboring and far-distance tissues and organs in multiple directions (**Figs 1-3**).

### Clinical application of cytocapsular tube-based precise cancer analyses in colon cancer

Next, we employed the precise cancer diagnosis with cytocapsular tube analyses in clinical applications. A man presented severe abdominal distention, pain, and constipation. He had no fevers, no chills, no hypothalamic amenorrhea or head injury, no shortness of breath, no wheezing, no hematuria, dysuria, no alerted level of consciousness, no dizziness, no new joint pains or muscle aches. He had no medical history of cancer. Computerized tomography scan examination revealed an obstructing colon cancer at the splenic flexure with secondary involvement of the greater curvature of the stomach.

He was performed an abdomen surgery. The surgery revealed that there is a colon tumor at the splenic flexure with 6.9 cm in length by 6.2 cm in circumference, grossly involving the omentum, serosa, and reaching into the portion of the attached stomach. A 19.3 cm colon in length with the tumor was resected. The lesion is 1.5 cm from the radial margin, 3.4 cm from the distal margin, and 8 cm from the proximal margin. The distal mucosa has normal rugae folds, and the proximal mucosa is focally attenuated but no additional lesions are noted. 17 lymph nodes are identified ranging from 0.2cm to 0.7cm in dimension with no positive lymph node as examined by microscope. There is a colon polyp, a single pink-tan tissue with 0.8 cm in the greatest dimension, at the proximal margin, which was identified as negative for high grade dysplasia.

H&E and immunohistochemistry staining (with antibodies MSH-2, MSH-6, MLH-1, and PMS-2) examination were performed with tissue specimens from the colon tumor block, paracancer tissues, 3.4 cm from the distal margin, 8 cm from the proximal margin, colon polyp, 17 lymph nodes, and resected stomach tissues. The pathology examination results are: colon cancer, adenocarcinoma, well to moderately differentiated, full thickness invasion with involvement of gastric serosa. Proximal margin, uninvolved; distal margin, uninvolved. 17 lymph nodes, 0 positive and margins are free of involvement. Polyp examined as tubular adenoma, negative for high grade dysplasia. Microscopic tumor extension: into serosa of stomach. Metastatic work up was negative so he had high risk stage IIC disease (high risk due to presentation of obstruction and invasion into the stomach). Pathologic staging: pT4bN0.

Cytocapsular tube (CT) analyses by immunohistochemistry staining and fluorescence microscope imaging were performed with tissue specimens from the same sites/slides in the above described samples. Cytocapsular tube results showed that there are many CTs and networks with variant CT densities in all the examined sites: the colon tumor niche, paracancer tissues, 3.4cm from the distal margin, 8cm from the proximal margin, colon polyp, 17 lymph nodes (attacking around lymph nodes but unable to concur yet), and resected stomach tissues (only at outer surface center NOT observed anywhere away from it or at the edge of the stomach tissue). Many colon cancer cells are in migration in the CTs and networks (**Fig. 4A-4G**). There are many CTs degraded into CT strand in diverse sizes (f **Fig. 4A-4G**). Cytocapsular tube-based Cancer Metastasis (CM)-grading: CM grade 4.

**Figure 4.**
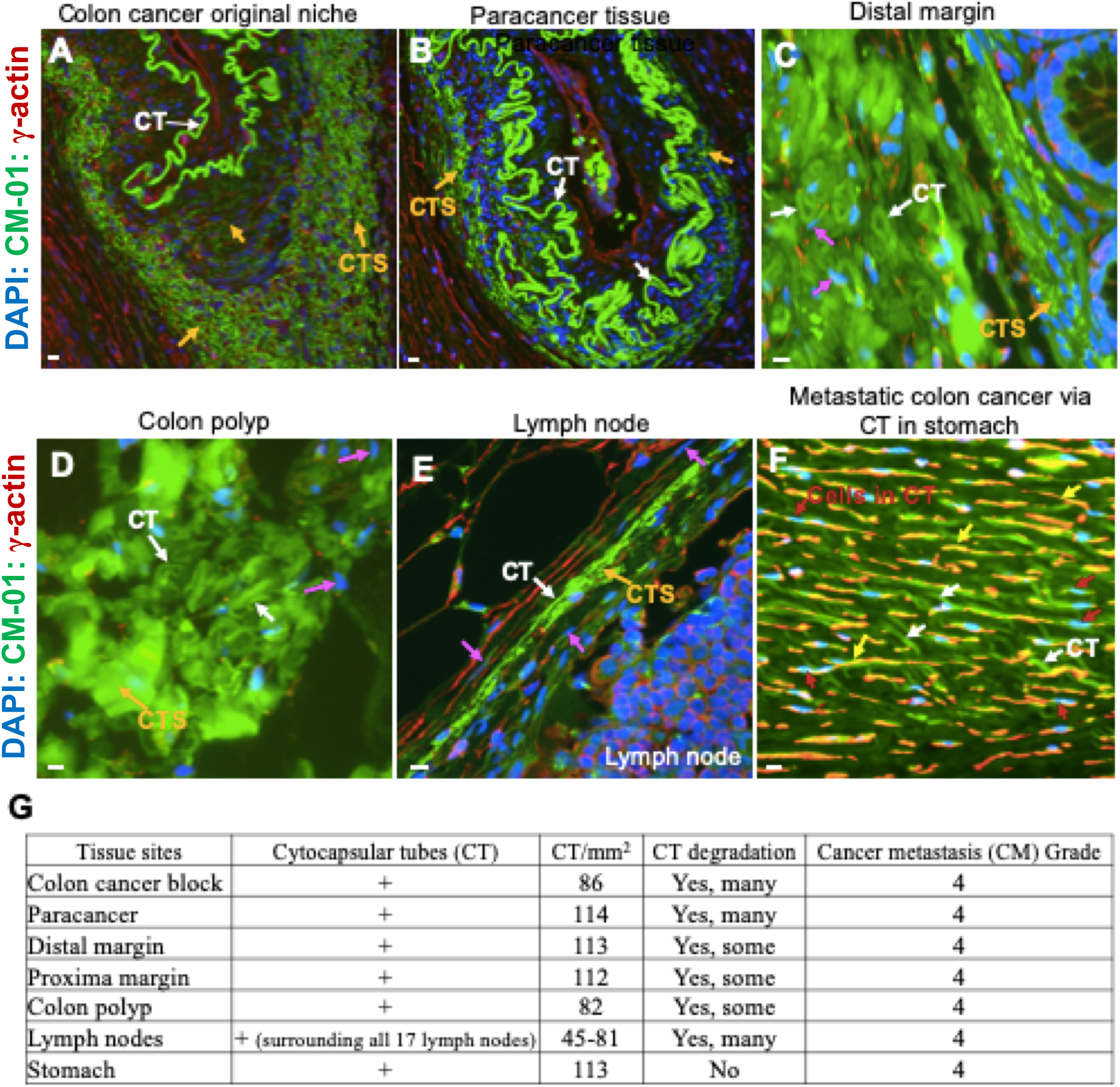
Clinical application of cytocapsular tube-based precise analyses in colon cancer diagnosis. **(A)** Fluorescence microscope image of colon cancer cell cytocapsular tubes (CTs, white arrows) in original niche. There are highly curved colon cancer cell CTs form super structures. Many colon cancer cell CTs degrade into thin CT strands (CTS, orange arrows). **(B)** Fluorescence microscope image of colon cancer cell cytocapsular tubes (CTs, white arrows) in paracancer tissues in the colon. There are some highly curved colon cancer cell CTs forming super structures. Some colon cancer cell CTs degrade into thin CT strands (CTS, orange arrows). **(C)** In the distal margin in the colon (8cm far from the colon tumor block), there are many CTs (white arrows) that form very dense CT masses. Colon cancer cells (purple arrows) are in migration in CTs. **(D)** In the colon polyps close to the colon cancer block, there are many CTs (white arrows) form CT masses, showing some CTs that degrade into very thin CT strands (CTS, orange arrow). Colon cancer cells (purple arrows) are in migration in CTs. **(E)** In the lymph node around the colon tumor block, there are some long CTs (white arrows) surrounding the lymph nodes with some CTs degradation into thick or thin CT strands (CTS, orange arrow). Colon cancer cells (purple arrows) are in migration in CTs. **(F)** In the stomach bottom tissue close to the colon cancer block, there are a large quantities of CTs (white arrows) in the stomach tissues. Many colon cancer cells (red arrows) are in migration in colon cancer cell CTs in stomach. The sectioned CTs are shown in golden color (yellow arrows) which are merged by green and red colors. **(G)** Quantitative analyses of CT densities for the discussed sites in this clinical application.

Because there were many cytocapsular tubes and colon cancer cells remaining in the colon, and a CM grade 4 assigned, we believed that the patient would benefit from adjuvant chemotherapy, likely with CapeOx. He took the chemotherapy of 4 cycles of CapeOx for 3 months and 1 month of 1cycle of Capecitabine alone. At 3, 6, 12, and 18 months after the chemotherapy completion, high contrast computer tomography scan, negative; carcinoembryonic antigen (CEA) test negative. At 2 years and 4 month after the surgical resection, he took a series of examinations: computer tomography scan, negative; ctDNA, negative, CEA, negative; four months and nineteen months past the surgery he had two colonoscopies done, all smooth colon except for a total of 3 benign polyps of 4-5 mm.

Overall, Cytocapsular tubes (CT, 3-6μm in diameter/width), CT networks and cancer cells within CT networks are invisible by current examination methods and technologies but may occur in the context of solid cancers, as is clearly seen in our patient’s tissue samples and may cause relapse. Therefore, cytocapsular tube analyses can provide an additional and precise analysis for evaluation of cancer cell metastasis in cancer prognosis and diagnosis, which may help to take proper treatments.

## Discussion

It is essential to understand the long-term elusive mechanics underlying colon cancer metastasis for the effective clinical diagnosis, therapy and treatment of colon cancers^1-4^. Here, we investigated colon cancer metastasis mechanics in 9 colon subtypes and evidenced that colon cancer cells generate membrane-enclosed physical freeway systems composed by cytocapsular tubes (CTs) and CT networks that conduct colon cancer cell metastases to both neighboring and far-distance tissues and organs.

Colon cancer cell cytocapsular tubes and the 3D CT networks form potent freeway systems for CT shielded and protected colon cancer cell dissemination in multiple directions. Colon cancer cell CTs are universally distributed in both compact and loose tissue of original niches, paracancer tissues and metastatic tissues and organs, displaying the powerful capacity of both individual CTs and collective CT masses in invading and passing through variable tissues with diverse textures and cell and ECM densities.

It is vital to precisely evaluate the colon cancer status in clinical colon cancer prognosis, before and after treatments (surgery, chemo-/radio-therapy and other treatments), and during colon cancer management to achieve better outcomes for patients^11-13^. Here, we applied the precise colon cancer diagnosis with CT analysis at the μm level in a clinical application. Colon is a long column-shaped organ and it is thought that colon cancer cells migrate and disseminate along the colon body with minimum/no multiple-direction metastases. According to conventional clinical colon cancer treatment protocols for Stage II colon cancer patients, it is thought that aggressive surgery and resection of the whole colon tumor blocks, and removing 4cm-8cm additional “normal” colons at both sides, would result in “complete eradication” of these colon cancers. Therefore, these patients do not need to follow up with chemotherapy as people thought that there is no colon cancer cells left and chemotherapy would be unnecessary. However, long-term clinical observations showed that for Stage II colon cancer patients undergone aggressive surgery but not followed up with chemotherapy, about 32% of the patients experienced colon cancer relapses within 6 months, and about 21% patients died within 1.5 years^14-15^. Clinical observations show that adjuvant chemotherapy for Stage II colon cancer is a clinical dilemma as the outcomes are debatable^15^. One possibility may be that the drugs have limited inhibition effects on CTs (off-target for CTs) and the colon cancer cells in the CTs, which protect colon cancer cells inside. We have shown that CT shielded cancer cells significantly and increase 10-100 fold the resistance to drugs that are off-targets to CTs (in another manuscript). Clinical oncology surgery, therapy and pathology doctors are always confused where the colon cancer cell come from after “eradication” by aggressive surgery^14-16^. Here, with the CT analyses, we clearly showed that even after aggressive colon-cancer surgery into the apparent “normal colon tissue” at both sides of the original niche, even 4cm to 8cm distance from the colon tumor blocks, there may still remain many CTs containing colon cancer cells. These remaining colon cancer cells in CTs would act as “seeds” for colon cancer relapse.

Here, this colon cancer patient obtained timely precise cancer diagnosis with CT analyses, and followed with an appropriate chemotherapy. After 2.3 years, with multiple kinds of the indicated examinations, there is no detectable sign of colon cancer relapse in the patient. Therefore, precise cancer diagnosis with cytocapsular tube analyses may provide an additional dimension for evaluation of cancer prognosis and diagnosis, which may help to take proper treatments without delay. The CTs in colon polyps suggests that precise CT-based cancer analysis may be applied in effective check/evaluation of colon polyps (benign or malignant), which are universally present in solid cancers with/without high cell density, and may be an effective method to fill the gap left by the high cell density based diagnosis methods (such as CT scan and PET-CT).

The colon cancer cytocapsular tube-based cancer analyses described here may facilitate the accurate evaluation of colon cancer status and metastasis, thereby improving therapy outcomes. To take advantage of the possibilities for the precise and CT-based colon cancer prognosis, diagnosis and evaluation of cancer status, particularly for very early stage colon cancer evaluation, more clinical analyses and applications are required. In the future, we expect that CT-based anti-colon cancer drug and technology development will significantly improve colon cancer pharmacotherapy and management.

## Supporting information

Ethics Waiver Letter

Ethics Statement

## Data Availability

All the data are included in the manuscript and available upon requests.

Supported by a grant (to Dr. Yi) from Cytocapsula Research Institute Fund for Cytocapsular Tube Conducted Cancer Metastasis Research, and a grant (to Dr. Yi) from Cellmig Biolabs Inc Fund for Cytocapsular Tube Cancer Metastasis Research.

Ethics Committee of Cellmig Biolabs Inc: Yubo Yang, PhD, Officer of Ethics Committee of Cellmig Biolabs Inc; Gerhard Wagner, PhD, Ethics Committee Member of Cellmig Biolabs Inc; Tingfang Yi, PhD, Ethics Committee Member of Cellmig Biolabs Inc; Herbert Lannon, PhD, Ethics Committee Member of Cellmig Biolabs Inc; Meghan Celli, Ethics Committee Member of Cellmig Biolabs Inc. Address: 245 First Street, Cambridge, MA, 02142 USA.

The study of this manuscript is based on the in vitro immunohistochemistry staining analyses with formalin -fixed and paraffin-embedded clinical colon cancer tissue slides, which were ordered from US Biomax (https://www.biomax.us; address: US Biomax, 15883 Crabbs Branch Way, Derwood, MD 20855, USA) or obtained from the Pathology Laboratory of Lake Forest Hospital of Northwestern University Medical School (address: 800 N Westmoreland Road, Suite 100, Lake Forest IL 60045 USA). All these tissue slides follow the rules and instructions of “Health Insurance Portability and Accountability Act”. Therefore, these assays do not require ethical oversight. Ethics Committee Member of Cellmig Biolabs Inc decided to waive the ethical oversight of this study.

We greatly acknowledge Dr. Ed Harlow, Harvard Medical School (USA) and Cancer Institute of University of Cambridge (UK), Dr. Jon Aster, Harvard Medical School, Dr. Jelle Wesseling, Leiden University (Netherlands), Dr. David Golan of Harvard Medical School (USA) for their help and meaningful discussion in the study.

