## Supplementary material for "Cytocapsular Tube-Based Precise Cancer Diagnosis in Colon Cancers": Ethics Waiver Letter

Cellmig Biolabs Inc.
245 First Street, Suite 1050
Cambridge, MA 02142

July 28, 2021

Dear MedRxiv Editor,

Cellmig Ethics Committee is in the charge of ethics related programs, procedures, supervision and management of all Cellmig and its subsidiaries, including Celldim Therapeutics Inc and Cytocapsula Research Institute (a non-profit research institute).

The study of the manuscript titled "Cytocapsular Tube-Based Precise Cancer Diagnosis in Colon Cancers" is based on the *in vitro* immunohistochemistry staining analyses with formalin -fixed and paraffin-embedded clinical colon cancer tissue slides, which were ordered from US Biomax (https://www.biomax.us; address: US Biomax, 15883 Crabbs Branch Way, Derwood, MD 20855, USA) or obtained from the Pathology Laboratory of Lake Forest Hospital of Northwestern University Medical School (address: 800 N Westmoreland Road, Suite 100, Lake Forest IL 60045 USA). All these tissue slides follow the rules and instructions of "Health Insurance Portability and Accountability Act". Therefore, these assays do not require ethical oversight.

Here, we decide and state that ethical oversight of this study is waived by the Ethics Committee of Cellmig Biolabs Inc.

Sincerely Yours,

Ethics Committee of
Cellmig Biolab Inc.

Signature: Yubo Yang
Name: Yubo Yang, PhD
Title: Officer of Ethics Committee
Cellmig Biolabs Inc.

Address:245 First Street, Suite 1050
Cambridge, MA 02142


Signature:
Name: Herbert Lannon, PhD
Title: Ethics Committee Member
Cellmig Biolabs Inc.

Address:245 First Street, Suite 1050
Cambridge, MA 02142
