## Supplementary material for "Cytocapsular Tube-Based Precise Cancer Diagnosis in Colon Cancers": Ethics Statement

Tingfang Yi, PhD
CEO & President
Cellmig Biolabs Inc.
245 First Street, Suite 1050
Cambridge, MA 02142
Cell:(617)785-7156


July 28, 2021

Dear MedRxiv Editor,

### Ethics Statement

The study of the manuscript titled "Cytocapsular Tube-Based Precise Cancer Diagnosis in Colon Cancers" is based on in vitro immunohistochemistry staining analyses of cytocapsular tubes with formalin-fixed and paraffin-embedded clinical colon cancer tissue slides.

The clinical application case in this study provide with in vitro cytocapsular tube analysis results for the post-surgery evaluation and diagnosis. The obtained results were interpreted by a qualified pathologist in conjunction with the patient's relevant clinical history, and other primary diagnostic tests, not an independent diagnosis application. Therefore, this study has no clinical trial ID.

These colon cancer tissue slides were ordered from US Biomax (https://www.biomax.us; address: US Biomax, 15883 Crabbs Branch Way, Derwood, MD 20855, USA) or obtained from the Pathology Laboratory of Lake Forest Hospital of Northwestern University Medical School (address: 800 N Westmoreland Road, Suite 100, Lake Forest IL 60045 USA). All these tissue slides follow the rules and instructions of "Health Insurance Portability and Accountability Act". Ethics Committee of Cellmig Biolabs Inc. decided to waive the ethical oversight of this study (detailed please see the Ethics Waiver Letter by Ethics Committee of Cellmig Biolabs Inc.).

Should you have any questions on ethics oversight or ethics statement, please never hesitate to contact me and we will respond you as soon as possible.

Sincerely Yours,

Tingfang yi

Tingfang Yi, PhD
